# Effect of isokinetic eccentric training on the human shoulder strength, flexibility, and muscle architecture in physically active men: a preliminary study

**DOI:** 10.1101/2023.10.15.23296999

**Authors:** Sebastian Vetter, Pierre Hepp, Axel Schleichardt, Stefan Schleifenbaum, Maren Witt, Christian Roth, Hans-Peter Köhler

**Author notes:** Corresponding author: (SV). These authors contributed equally to this work (Christian Roth, Hans-Peter Köhler).

## Abstract

Strengthening the rotator cuff muscles is important for injury prevention and rehabilitation. However, there is considerable evidence that range of motion and fascicle length are risk factors for injury. Since muscle fascicle length improves motor performance and reduces the risk of injury, it may be an important variable to promote multidirectional changes in the function and macroscopic structure of the target muscle. As a well-established intervention strategy, passive stretching seems best for improving the flexibility in terms of range of motion. But stretching and range of motion seem to have little effect on fascicle length changes and injury risk. However, recent reviews suggest that eccentric exercises improve functional and structural measures for the lower limbs. Since a research gap exists for the shoulder joint it has to be clarified if eccentric training for the external rotator cuff muscles produces similar functional and structural changes for a non-specific shoulder trained healthy population. The purpose of this study was to evaluate the effects of highly standardized eccentric isokinetic resistance training on the function and structure of the dominant shoulder external rotator cuff muscles. In addition, the feasibility of a new method of measuring flexibility and muscle diffusion tensor imaging to quantify three-dimensional changes in muscle fibers was tested. Therefore, 16 physically active men were recruited in October 2021. The right shoulder eccentric strength was trained twice a week for six weeks at 30°/s using an isokinetic dynamometer. The eccentric training protocol consisted of five sets and ten repetitions with subjective maximal effort. The primary outcome measures were for the external rotator cuff muscles eccentric and concentric mean and peak torque at 30, 60 and 180°/s, flexibility in terms of active and passive stretching, fascicle length and fascicle volume for the supraspinatus and infraspinatus muscles. The findings showed a training effect for eccentric strength (+24%). For the external rotator cuff muscles, the torque-angle relationship increased, especially in the final phase of range of motion, even though a 4% decrease was seen after the passive stretch test. Positive changes in muscle structure were shown for fascicle length (+16%) and fascicle volume (+19%). Secondary outcome measures for the internal rotator cuff muscles did not show significant changes from baseline to post-test. Based on the study results, we can conclude that eccentric isokinetic training has a significant effect on the function and macroscopic structure of the external rotator cuff muscles in a non-specifically trained, healthy male population. To our knowledge, this is the first eccentric training study using both isokinetic dynamometer and muscle diffusion tensor imaging to access functional and structural changes in the human shoulder rotator cuff muscles. The methods were shown to be feasible for interventional studies. Based on these results, populations such as high-performance handball players with highly trained shoulders should be included in future studies.

## Introduction

Shoulder injuries are an ongoing problem in general practice[1]. The lifetime prevalence of shoulder problems is approximately 31 %[2]. Key factors for the reduction of injury rates seem to be joint range of motion (ROM) and strength[3–6]. Structural deficits in muscle fascicle length (FL), which are also associated to ROM and strength, can also be seen as potential risk factors[7,8]. However, most exercise-based interventions appear to target changes in functional outcomes for flexibility and strength without significant reduction in injury rates[9–11].

In contrast, a recent meta-analysis of 8459 athletes shows that eccentric field exercises implemented in sports athletic training halve the risk of hamstring injuries[12]. Eccentric training effectively induces tissue damage and hypertrophic response. Interestingly, following eccentric strength training, fiber lengthening and a decrease in fiber pennation angle seem to be a major reason for a positive change in muscle volume and motor performance[13]. However, exercises that promote muscle fascicle lengthening seem to be of considerable interest as injury prevention strategies and for improving motor performance[13–15].

Recently, eccentric training interventions have received greater attention in research with the objective of better understanding functional and structural changes through these forms of interventions[16,17]. However, there is still a lack of studies focusing on the musculature of the shoulder joint[18]. Furthermore, since structural changes are commonly obtained with 2D ultrasound imaging, even less is known about the three-dimensional changes in muscle fiber architecture. As a promising approach muscle diffusion tensor imaging (mDTI) is able to show the complexity of changes in different muscles[19–21].

The aim of this study was to investigate the structural and functional alteration following a six-week isokinetic eccentric strength training. In addition, this study will test the feasibility of a newly developed isokinetic functional stretch test as well as the feasibility of mDTI to quantify changes in the muscle parameters for the shoulder external rotator cuff muscles. Main outcome measures were eccentric and concentric mean and peak torque of the external rotator cuff muscles at an isokinetic mode with 30°/s, 60°/s, and 180°/s movement velocity and analysis of the torque angle relationship. For flexibility, active ROM (aROMmax) and submaximal and maximum passive ROM (pROMsub and pROMmax) and its passive torque angle relation were obtained. To quantify changes in the infraspinatus and supraspinatus muscle, mean fractional anisotropy (FA), fascicle length (FL) and fascicle volume (FV) were calculated based on mDTI.

## Materials and Methods

This is a preliminary experimental study[22]. The recruitment period for this study was from October 18 to December 17, 2021. The study protocol was approved by the Leipzig University ethics committee (ethical approval nr: 362/21-ek) and is registered in the German Clinical Trials Register (DRKS00032375). The study was performed according to the declaration of Helsinki. All subjects gave written informed consent. The authors had no access to information that could identify individual participants during or after data collection.

### Sample

The sample consisted of 16 active male student athletes for isokinetic measurements (23.3 ± 3.9 years; 181.2 ± 6.9 cm body height; 76.4 ± 7.4 kg body mass). The physical activity of the recruited population is characterized by strength training and individually varied activities such as skiing, judo, soccer, running, handball, or javelin throwing at least twice a week. Out of this population eleven subjects were randomly chosen for magnetic resonance imaging (MRI). All participants were required to have a healthy dominant right shoulder. Subjects were excluded, if they showed a history of musculoskeletal disorders, had regular medication, discomfort, pain, or known lesions in the right shoulder. In addition, five days prior to data collection, participants were instructed to discontinue any intense and exceptional physical activity or heavy training.

### Eccentric isokinetic resistance training intervention

A total of twelve eccentric isokinetic strength training sessions for the external rotator cuff muscles were performed over a period of six weeks (twice a week). Training was performed using BTE Primus RS isokinetic dynamometer (Baltimore Therapeutic Equipment Company, Hanover, MD, USA). Prior to the start of the intervention, all subjects completed two sessions of familiarization with the eccentric training and testing procedure.

The training protocol followed the recommendations from Toigo and Boutellier[23] and was standardized in terms of weeks and days of training, ROM, movement velocity, load magnitude, number of sets and repetitions, and exercise execution regulations (Table 1). After two weeks of intervention, the total ROM was shifted 15 degrees toward the internal ROM and the load level was increased based on the load level of the last three sessions. The training was performed in prone position with 90° shoulder abduction and 90° elbow flexion.

**Table 1.**
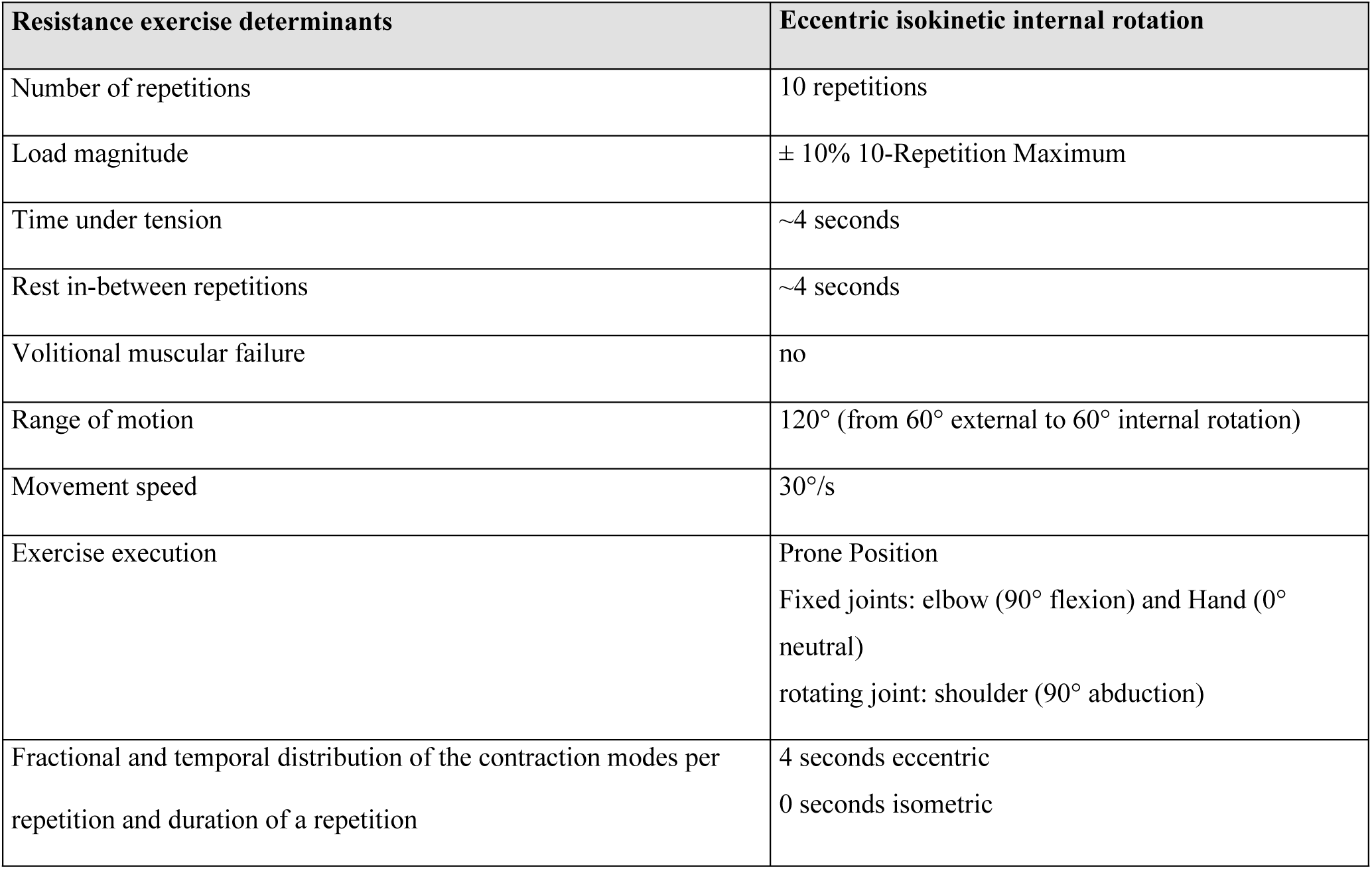
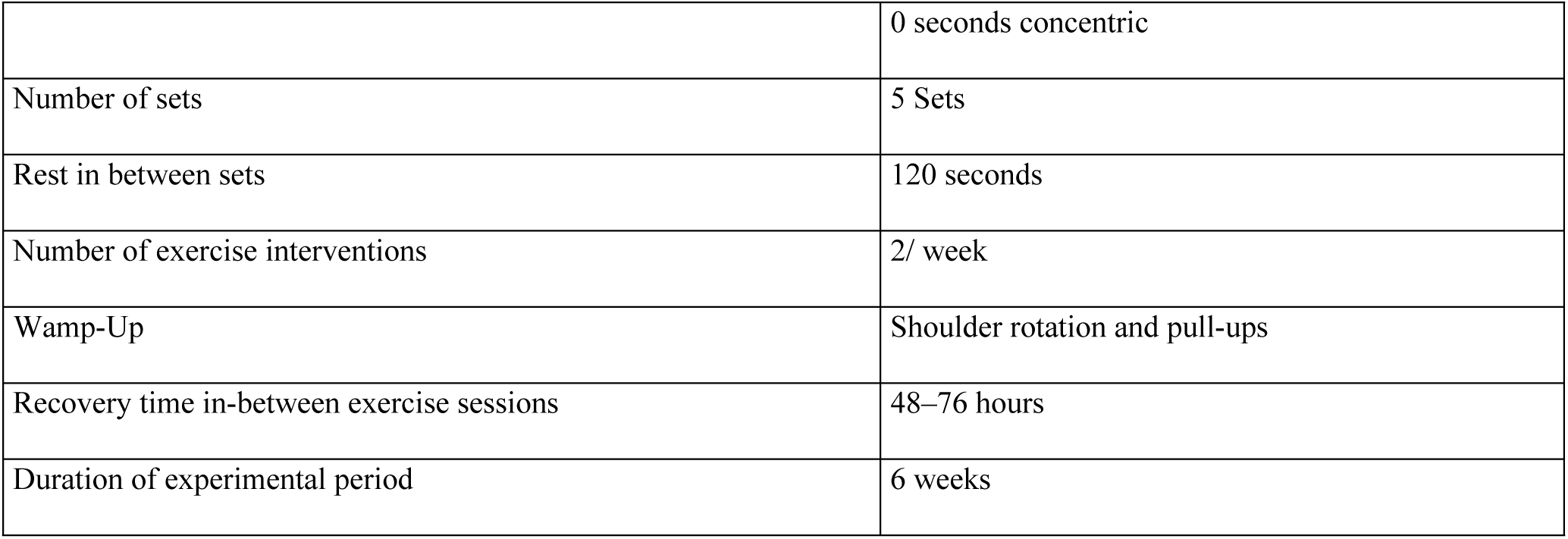
Eccentric resistance exercise determinants.

| Resistance exercise determinants | Eccentric isokinetic internal rotation |
| --- | --- |
| Number of repetitions | 10 repetitions |
| Load magnitude | ± 10% 10-Repetition Maximum |
| Time under tension | ~4 seconds |
| Rest in-between repetitions | ~4 seconds |
| Volitional muscular failure | no |
| Range of motion | 120° (from 60° external to 60° internal rotation) |
| Movement speed | 30°/s |
| Exercise execution | Prone Position<br>Fixed joints: elbow (90° flexion) and Hand (0° neutral)<br>rotating joint: shoulder (90° abduction) |
| Fractional and temporal distribution of the contraction modes per repetition and duration of a repetition | 4 seconds eccentric<br>0 seconds isometric |
|  | 0 seconds concentric |
| Number of sets | 5 Sets |
| Rest in between sets | 120 seconds |
| Number of exercise interventions | 2/ week |
| Wamp-Up | Shoulder rotation and pull-ups |
| Recovery time in-between exercise sessions | 48–76 hours |
| Duration of experimental period | 6 weeks |

### Data acquisition

Functional and structural parameters of the right shoulder were assessed using diffusion-weighted MRI scans and isokinetic dynamometry three to five days before the first and after the final eccentric intervention session.

### Isokinetic Testing

IsoMed 2000 (D&R Ferstl GmbH, Hemau, Germany) was used to test the ROM and strength of the right shoulders external and internal rotator cuff muscles. Each subject was tested in the supine position with fixed shoulder-arm joints (Fig 1) as for the eccentric training described (Table 1). In addition, the shoulder was fixed ventrally to prevent shoulder elevation in an individually standardized manner.

**Fig 1.** Functional testing using ISOMED 2000 isokinetic dynamometer.

Active stretch testing consisted of 15 trials alternating between internal and external rotation. Volunteers were instructed to move the apparatus slowly and with ease until aROMmax. The turning point were reached when the subject was unable to continue the antagonist stretch generated by the agonist muscles. Passive stretching tests were performed at an isokinetic speed of 5°/s. The passive stretch involved 15 trials of alternating internal and external rotation with eyes closed and muscles subjectively relaxed until pROMmax, characterized by 8 Nm of passive torque, was reached.

The voluntary maximum strength tests were performed separately for concentric and eccentric rotation, also alternating between internal and external rotation. Strength measurements were performed over a 160° ROM (70° internal rotation and 90° external rotation) at an isokinetic speed of 60/°s, 180°/s and 30°/s. All tests were performed with a rest period of 60 seconds. Each subject’s position and dynamometer adapter gravity correction value and settings were individualized at baseline and used for post-tests (Fig 1).

### Magnetic Resonance Imaging

A 3-Tesla Siemens MAGNETOM Prisma scanner (Siemens Healthcare, Erlangen, Germany) with a shoulder coil (XL, 16-channel) was used for MRI scans. The participants lay in a head first supine position with the right arm in the neutral position and the hand supinated. The MR protocol consisted of a T1-weighted and a diffusion-weighted sequence. The T1-weighted sequence consisted the following parameter settings: repetition and echo time TR/TE = 492/20 ms, slice thickness = 0.7 mm, flip angle = 120°, field of view = 180 x 180 mm^2^, matrix = 256×256 mm^2^. Diffusion-weighted sequence were acquired as following: repetition and echo time TR/TE = 6100/69 ms, slice thickness = 5.2 mm, flip angle = 90°, field of view = 240 x 240 mm^2^, matrix = 122 x 122 mm^2^, 48 diffusion sampling directions with b = 400 s/mm^2^. Total scan time reached 12 minutes.

### Data processing

MATLAB v.R2022a (MathWorks, Natick, USA) was used to process the isokinetic dynamometer data. Stretching parameter calculations were based on the average of the third through seventh trials in each test, with the first two trials excluded and defined as familiarization. pROMmax was calculated when subjects reached the end point characterized by either 8 Nm or 100° internal rotation and 130° external rotation depending on the target muscle for the test. For further analysis of changes in the morphology of the passive torque-angle curves, a three-parametric e-function (Appendix) was fitted from each subject’s individual offset start angle to the 100° internal rotation and 130° external rotation end points (100% stretch), which are shown in percent stretch in the figures. The evaluated e-function was then used to calculate the angle at the instant of 0.01 Nm/degree (pROMsub) and to calculate the fit to the defined end points using a statistical parametric mapping (SPM1d) method in MATLAB (Appendix).

For strength analysis, the mean of three maximum consecutive repetitions were used. Raw data were filtered using a 6 Hz cutoff frequency[24]. From each test, the acceleration and deceleration phases were cut, leaving only the interval with the desired isokinetic velocity. The maximum torque was then identified in the isokinetic phase and the mean torque was calculated over this phase. Descriptive and inferential statistics in SPSS v.27 (IBM. Armonk. New York. USA) used absolute torque. For comparison of curve morphology using SPM1d, data were normalized to body mass[24,25].

MRI data were first processed using Mimics Materialise v.24.0 (Leuven, Belgium) for manually segment the supraspinatus and infraspinatus muscles to extract a volume of interest (VOI). This segmentation method showed excellent reliability for the tractography outcome measures[26]. Then, the diffusion-weighted images were processed and corrected using DSI Studio (v. 3rd of December 2021. http://dsi-studio.labsolver.org). Based on VOI-based deterministic fiber tractography, FA, muscle FL and FV were calculated using the integrated statistics tool in DSI Studio (Fig 2). More details for the MRI and segmentation can be found in a previous methods paper[26].

**Fig 2.**
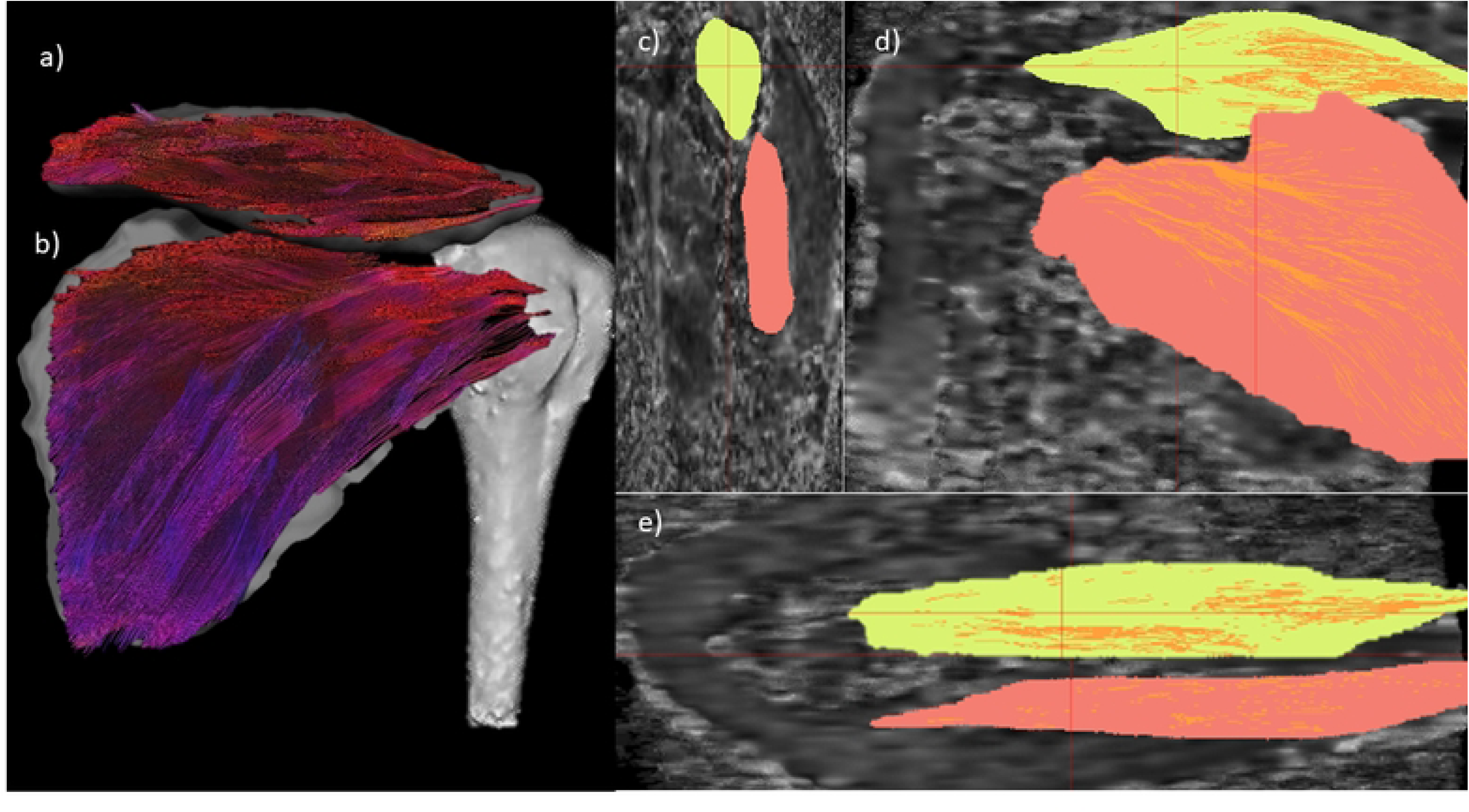
Right shoulder dorsal view tractography. a) coronal plane supraspinatus muscle fiber with directional coloring, b) infraspinatus muscle, c-e) sagittal, coronal and axial view on diffusion-weighted-images and volume of interest.

### Statistics

MATLAB v.R2022a (MathWorks, Natick, USA) and SPSS v.27 (IBM, Armonk, New York, USA) were used for statistics. Descriptive results were based on mean and standard deviation (±). Participants were excluded from further analysis if the data distribution showed outliers and z-transformed values exceeded 2.5. For statistical analysis, missing variables were calculated in SPSS. Repeated measures multivariate analysis of variance (MANOVA) was used to show main effects and interactions between different factors. For statistics of strength parameters, the factors were time (pre and post), mode (eccentric and concentric), and speed (30°/s, 60°/s and 180°/s). For flexibility parameters, the factors were time (pre and post) and test (aROM and pROM). For muscle parameters, the factor for statistical analysis were time (pre and post). Statistics were separated for the muscles.

For further post-hoc mean comparisons, repeated measures univariate analysis of variances (ANOVA) and paired t-tests were used. The *p*-value was set at 0.05. Differences between the torque-angle curves were calculated using SPM1d integrated in MATLAB.

## Results

All 16 subjects completed twelve training sessions within six weeks. Data were tested for normal distribution and homogeneity. One outlier was excluded for supraspinatus muscle analysis, and three subjects were excluded due to invalid test performance for the active stretch test and a truncated proximal field of view on MRI for the infraspinatus muscle.

### External rotator cuff muscles functional and structural changes

A three-factors repeated measures MANOVA were calculated for external and internal rotator cuff muscles strength. Overall, the external rotator cuff muscles showed a significant time x mode interaction for the parameter mean torque (F (1,14) = 5.82; *p* = .030; ηp^2^ = .293) and peak torque (F (1,14) = 3.58; *p* = .080; ηp^2^ = .203). Also, an interaction effect was found for time x speed for mean torque: (F (1,14) = 5.29; *p* = .011; ηp^2^ = .274) and peak torque (F (1,429; 20,004) = 3.51; *p* = .063; ηp^2^ = .200). The interaction effect of time x mode was significant for the 30°/s velocity strength tests (Wilks-Lambda = .450; F (2,13) = 7.93; *p* = .006; ηp^2^ = .550).

For analysis of the stretching tests and ROM results, a one-way MANOVA showed significant pre-post decreases for the trained external rotator cuff muscle parameters (Wilks-Lambda = .572; F (2,14) = 5.25; *p* = .020; ηp^2^ = .428). Furthermore, the analysis for the aROMmax revealed a significant pre-post decrease for the internal rotator cuff muscles (t (12) = 2.37; *p* = .018; d = .657). The pre-post differences for the external rotator cuff muscles for each factor and parameter can be found in Table 2.

**Table 2.** Results for the external rotator cuff muscles.

| Muscle | parameter | | Pre | Post | $\Delta\%$ | $p$ |
| --- | --- | --- | --- | --- | --- | --- |
| External rotator cuff muscles | Strength eccentric (Nm) |  |  |  |  |  |
|  | Mean torque | 30°/s | 28.68 (6.31) | 35.57 (6.58) | +24.02 | .008 |
|  |  | 60°/s | 33.01 (6.92) | 37.42 (7.31) | +13.36 | .098 |
|  |  | 180°/s | 36.60 (7.97) | 40.73 (8.42) | +11.30 | .145 |
|  | Peak torque | 30°/s | 37.70 (7.76) | 44.04 (7.80) | +16.82 | .031 |
|  |  | 60°/s | 41.12 (8.25) | 45.34 (8.93) | +10.25 | .150 |
|  |  | 180°/s | 42.50 (8.94) | 46.22 (9.29) | +8.76 | .190 |
|  | Strength concentric (Nm) |  |  |  |  |  |
|  | Mean torque | 30°/s | 22.55 (4.52) | 23.07 (4.10) | +2.32 | .451 |
|  |  | 60°/s | 22.86 (4.23) | 24.45 (4.57) | +6.93 | .227 |
|  |  | 180°/s | 21.75 (4.55) | 21.48 (4.14) | -1.29 | .374 |
|  | Peak torque | 30°/s | 28.61 (5.54) | 30.49 (5.41) | +6.57 | .226 |
|  |  | 60°/s | 29.41 (5.25) | 31.38 (6.31) | +6.70 | .253 |
|  |  | 180°/s | 25.69 (4.63) | 25.23 (4.77) | -1.81 | .345 |
|  | Flexibility (internal rotation) |  |  |  |  |  |
|  | Active stretch | aROMmax (°) | 71.59 (8.62) | 66.82 (7.72) | -6.66 | .018 |
|  | Passive stretch | pROMsub (°) | 60.95 (10.70) | 58.06 (9.48) | -4.74 | .009 |
|  |  | pROMmax (°) | 86.63 (9.65) | 83.46 (8.50) | -3.66 | .002 |
| Infraspinatus | Fascicle length (mm) |  | 66.55 (6.15) | 64.63 (4.80) | -2.89 | .277 |
|  | fascicle volume (mm<sup>3</sup>) |  | 257.83 (30.21) | 258.29 (35.33) | +0.18 | .492 |
|  | Fractional anisotropy |  | 0.2500 (0.02) | 0.2486 (0.02) | -0.56 | .690 |
| Supraspinatus | Fascicle length (mm) |  | 36.76 (6.93) | 42.68 (7.94) | +16.1 | .003 |
|  | fascicle volume (mm<sup>3</sup>) |  | 143.17 (29.34) | 170.54 (23.42) | +19.12 | .002 |
|  | Fractional anisotropy |  | 0.2855 (0.02) | 0.2845 (0.02) | -0.35 | .889 |

Columns indicate the absolute mean pre and post test values, percentage change ( $\Delta\%$ ) and significance ( $p$ ). Nm, newton meter; aROMmax, maximum active range of motion; pROMsub, submaximal passive range of motion; pROMmax, maximum passive range of motion; °, angle degree; °/s, degrees per second (movement velocity); mm, millimeter; mm<sup>3</sup>, cubic millimeter; values in brackets, standard deviation.

Muscle structural changes were calculated for the supraspinatus muscle and showed a non-significant result (Wilks-Lambda = .363; F (2;8) = 7.03; *p* = .017; ηp^2^ = .637). However, post-hoc tests for the pre-post differences can be found for FL (t (9) = 3.53; *p* = .003; d = 1.117) and FV (t (9) = 3.82; *p* = .002; d = 1.207). The Infraspinatus muscle did not show statistically significant pre-post differences (Table 2). Furthermore, no changes were found for the metric FA and for all secondary outcome measures (Table 3).

**Table 3.**
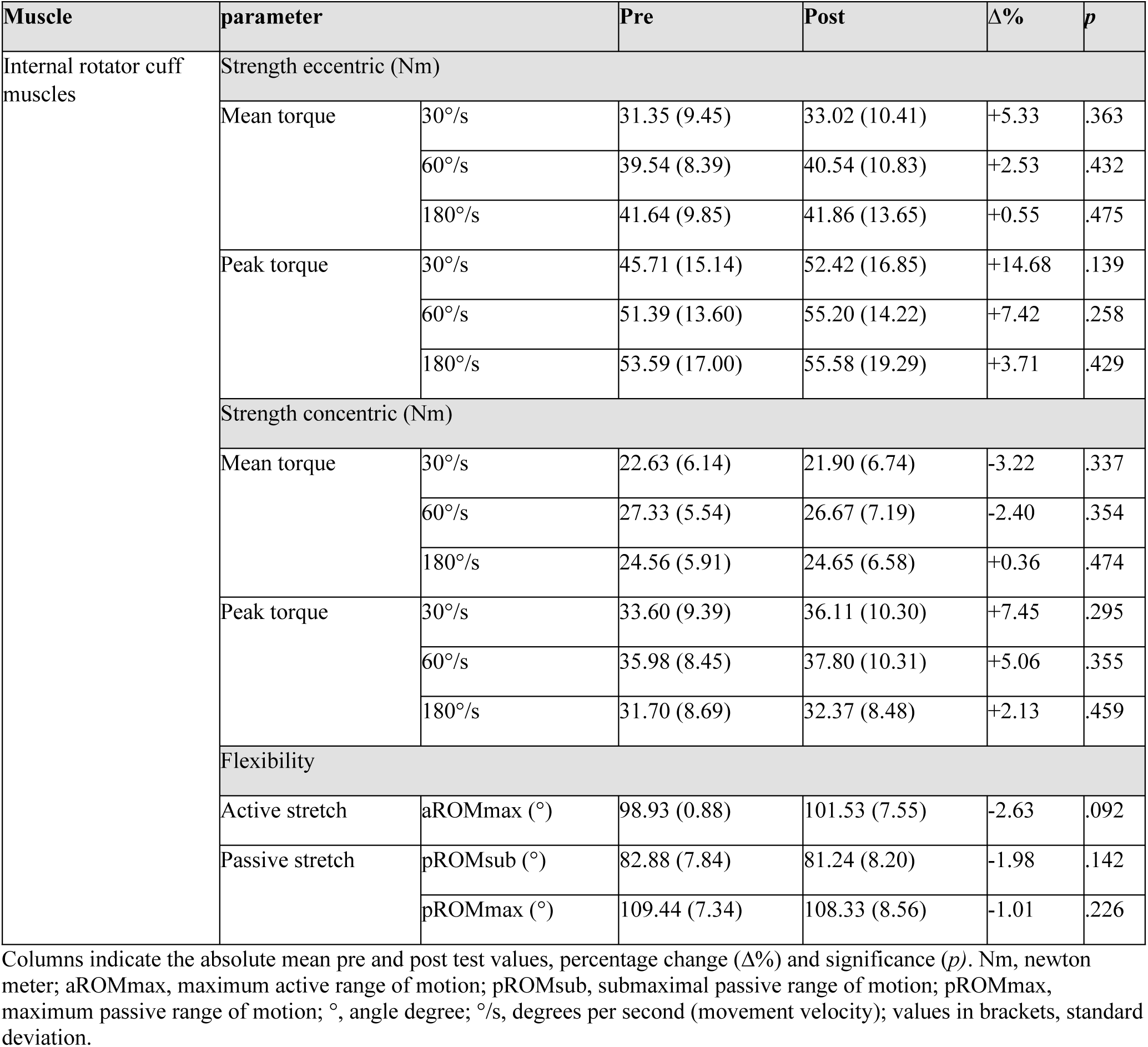
Results for the internal rotator cuff muscles.

### Internal rotator cuff muscles functional changes

A three-factors repeated measures MANOVA were calculated for the internal rotator cuff muscles strength showing no significant changes in all metrics Furthermore, a one-way MANOVA for the stretching tests showed no significant changes (Table 3).

### Changes in eccentric and concentric torque-angle curves

For the external rotator cuff muscles strength tests, SPM1d analysis of the torque-angle relationship revealed significant changes between neutral position and 60° internal rotation in the eccentric test with an isokinetic speed of 30°/s. The eccentric test with 60°/s movement velocity showed a significant increase in torque between 30 and 60° internal rotation(Fig 3). SPM1d analysis of the internal rotator cuff muscles showed a significant change in curve morphology in the internal rotation in the 60-40° ROM and in the final ROM above 80° (Fig 4).

**Fig 3.**
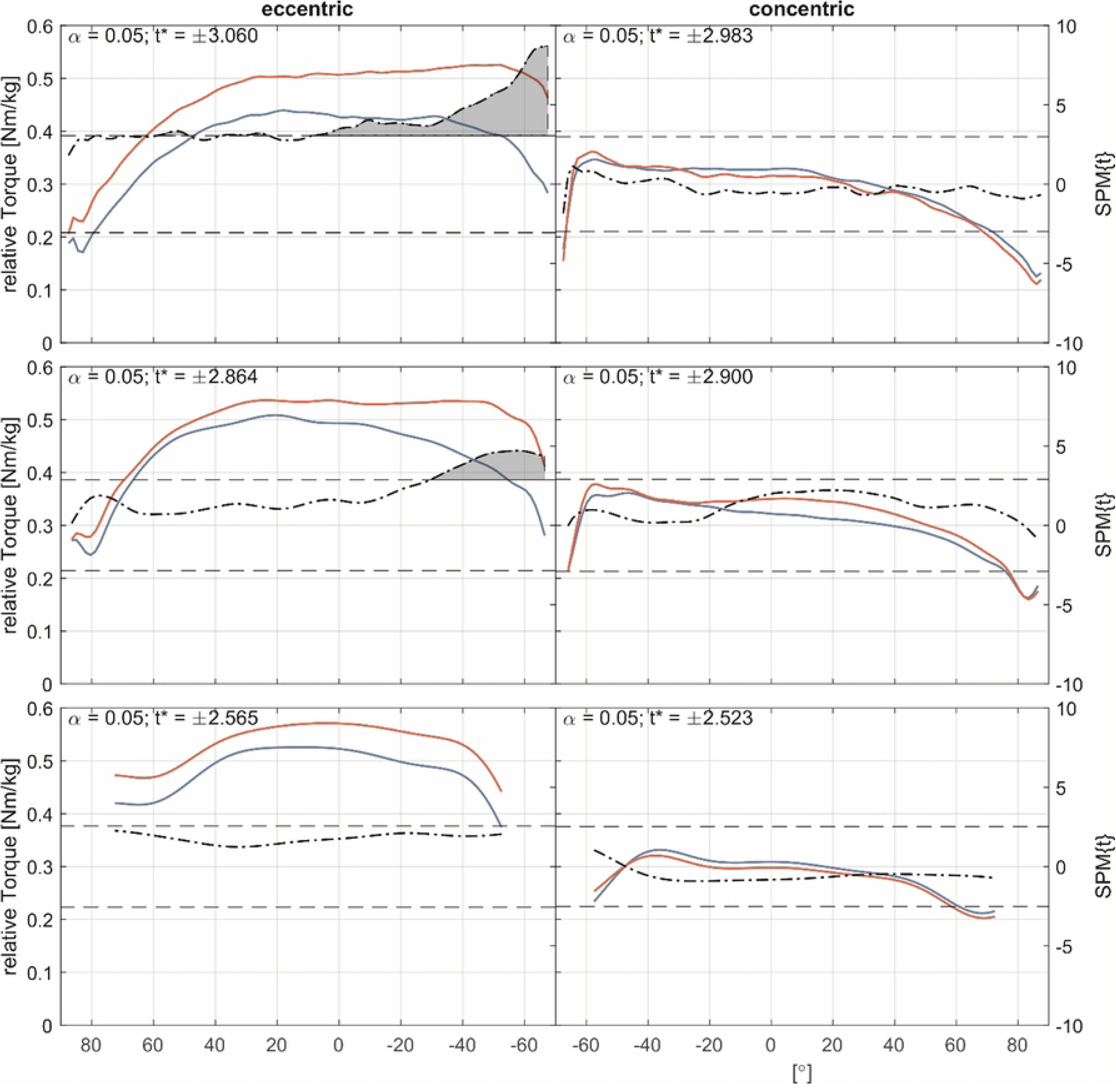
External rotator cuff muscles torque-angle-curves. Pre- (blue) and Post-measurement (red) torque angle curves for 30°/s, 60°/s and 180°/s (top-to-bottom) for the eccentric (left column) and concentric (right column) strength. The dash-dotted line indicates the *t*-values calculated by the Statistical Parametric Mapping method (SPM1d) for each angle, the dashed line indicates the critical *t*-value. If the *t*-values exceed the critical value, significant differences exist between pre- and post-measurement and are marked as grey area.

**Fig 4.**
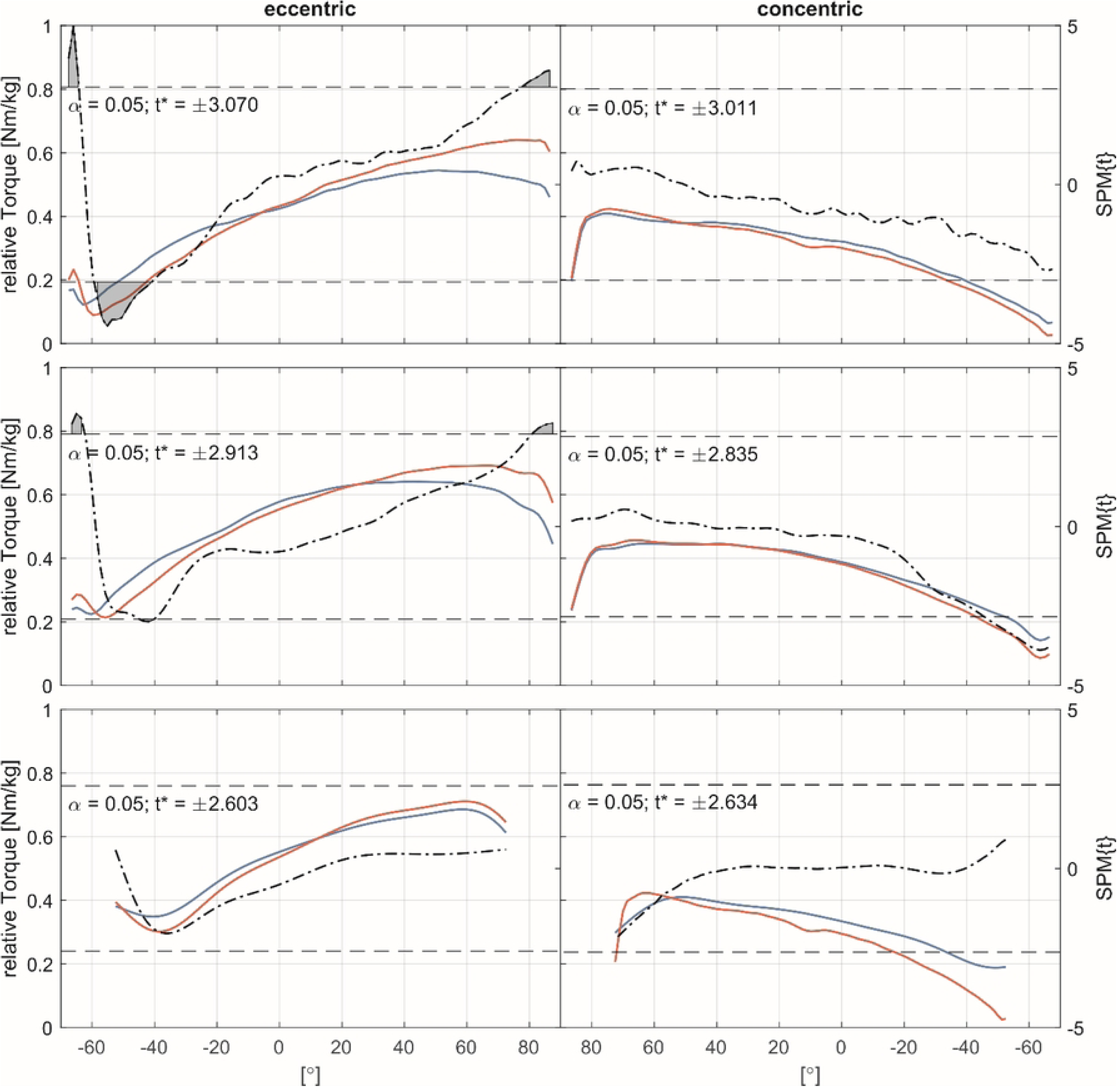
Internal rotator cuff muscles torque-angle-curves. Pre- (blue) and Post-measurement (red) torque angle curves for 30°/s, 60°/s and 180°/s (top-to-bottom) for the eccentric (left column) and concentric (right column) strength. The dash-dotted line indicates the *t*-values calculated by the Statistical Parametric Mapping method (SPM1d) for each angle, the dashed line indicates the critical *t*-value. If the *t*-values exceed the critical value, significant differences exist between pre- and post-measurement and are marked as grey area.

### Changes in passive torque-angle curves

For the external rotator cuff muscles passive stretching tests SPM1d analysis showed an increase in passive stretching torque-angle relationship from 40° ROM to 100° ROM (Fig 5). No differences in passive torque-angle relationship were statistically confirmed for the internal rotator cuff muscles (Fig 6 and Table 3).

**Fig 5.**
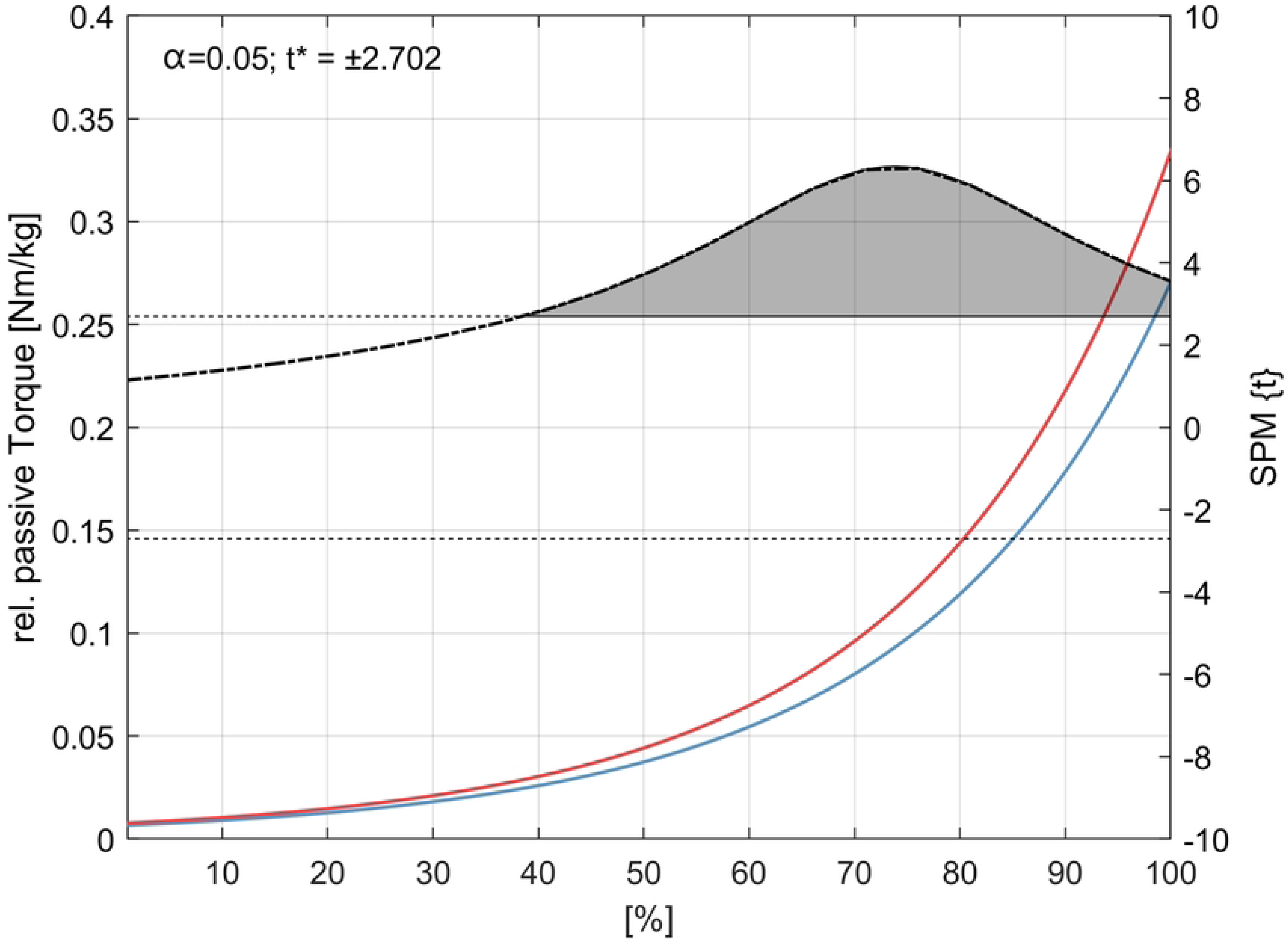
External rotator cuff muscles passive stretching torque-angle-curves. Pre and post group torque angle curves (blue and red) extrapolated from the individual offset start angle of each subject to 100° of internal rotation (100% extension during testing). Newton meters (Nm) are expressed relative to the body mass (kg) of each subject. The dotted line indicates the *t*-values calculated by the statistical parametric mapping method (SPM1d) for each angle, the dashed line indicates the critical *t*-value. Significant differences are indicated by the gray area in the figure.

**Fig 6.**
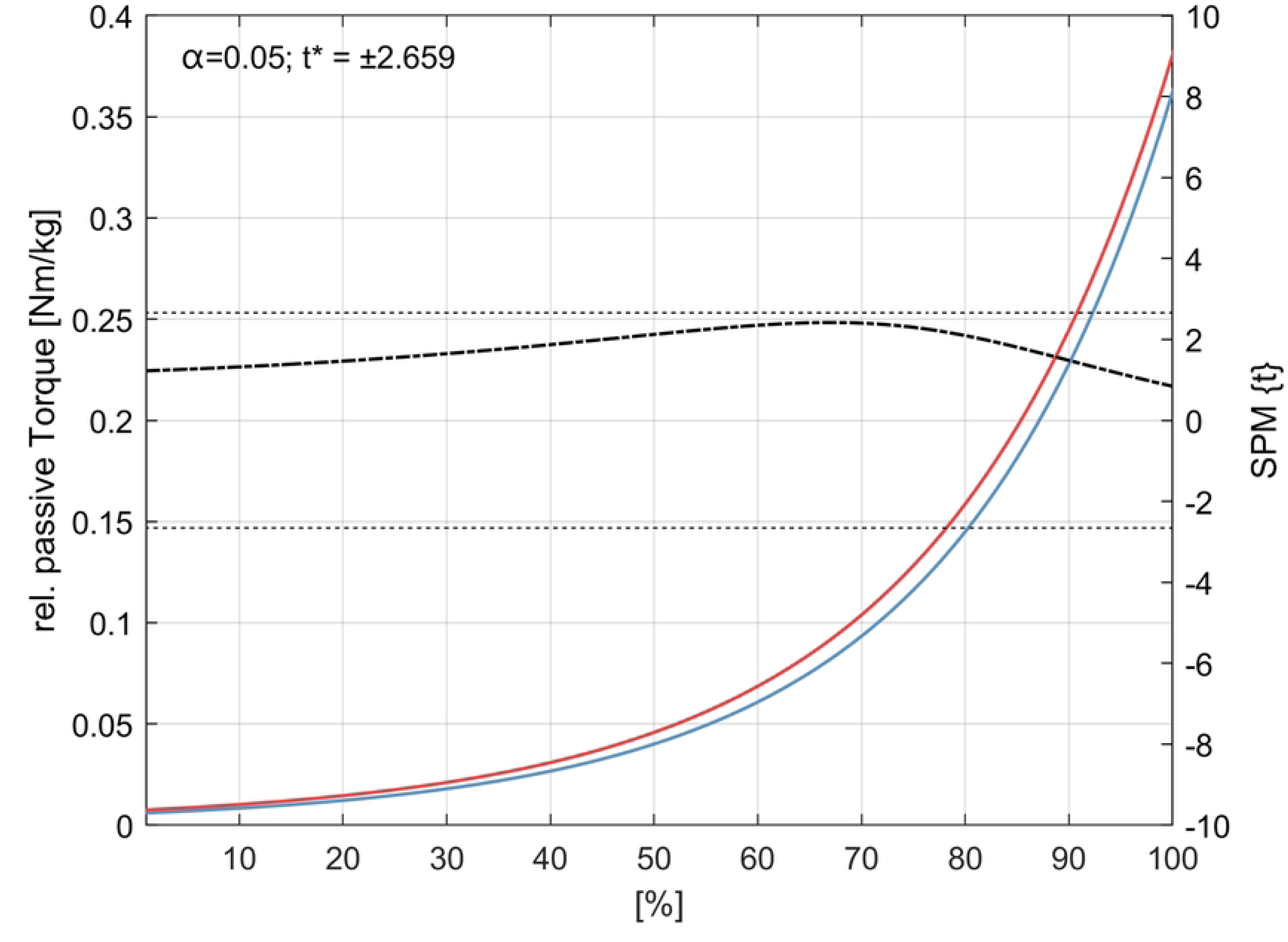
Internal rotator cuff muscles passive stretching torque-angle-curves. Pre-post (blue-red) group torque-angle curves extrapolated from each subject’s individual offset start angle to 130° of external rotation (100% extension in test). Newton meters (Nm) are relative to each subject’s body mass (kg). The dash-dotted line indicates the *t*-values calculated by the Statistical Parametric Mapping method (SPM1d) for each angle, the dashed line indicates the critical *t*-value. Significant differences are indicated by the gray area.

## Discussion

The purpose of this study was to determine if eccentric resistance training could produce similar and plausible results for the shoulder compared to other lower extremity studies and to evaluate the feasibility of the study methodology. We hypothesized that 6 weeks of 30°/s isokinetic eccentric internal rotation resistance training targeting the external rotator cuff muscles, would have positive effects on eccentric strength, active and passive stretching behavior, and supraspinatus and infraspinatus muscles FA, FL and FV. However, the eccentric strength of the external rotator cuff muscles increased significantly by +24% in the 30°/s test in particular in the final ROM of the isokinetic phase (Fig 3). Flexibility tests showed a significant decrease in both the aROMmax (-7%) and the pROMmax (-5%), while the passive torque angle relation increased. In addition, the supraspinatus FL increased significantly by +16% and the FV increased by +19%. Other primary outcome measures, such as concentric strength for the trained external rotator cuff muscles, changed up to +7% but not statistically significant. Secondary outcome measures showed no statistically significant changes in internal rotator cuff strength and flexibility tests.

Strength tests results are plausible and comparable to other lower extremity eccentric isokinetic dynamometer training studies, explaining a 19% gain in eccentric strength and a 9% improvement for the concentric strength[18]. Several mechanisms and adaptations may account for the changes in strength. First, the changes in strength and FV are addressed in terms of calculated fiber elongation[13] and the documented right shift in the torque-angle relationship (Fig 3), which has been discussed as a functional correlate of sarcomere formation[13,23,27]. Second, the changes found are primarily concerned with possible changes in FV as a possible surrogate parameter for physiological cross-sectional area of a muscle.

As multiple tissues define the ROM of the shoulder, a decrease in flexibility can be attributed to the function and structure of the glenohumeral joint. Since the shoulder joint consists of at least three other joints, not only the stabilizing muscles, but also passive structures such as the capsule determine its function. Therefore, Zandt et al.[28] expected low adaptability for rotator cuff muscles following eccentric resistance training. Furthermore, Camargo and collegues[29], summarized that the stimulation of the target muscle seems to be problematic for shoulder rotational movements. However, this study showed interesting changes in joints flexibility while parametrizing the passive stretching torque-angle curves and extracting parameters that characterize the changes in the stretching behavior until the final stretch was reached. Our results show that a decrease in ROM after eccentric resistance training can be explained by an increase in the torque-angle relationship during the passive stretch test. This can be explained by changes in stiffness either due to addition in sarcomeres in series or due to chronic titin elongation and therefore sarcomere lengthening[30,31]. Therefore, this approach provides new information on the stretch behavior of human shoulder external rotator cuff muscles after resistance training without using imaging techniques to quantify tissue stiffness[32].

This study shows a 16% increase in FL following eccentric training. Changes in FL can be attributed to either sarcomere elongation or sarcomerogenesis[30,31]. However, this study found a positive change in FV and in torque-angle-relationship, which can be interpreted as an indicative for sarcomerogenesis, as these changes appear to be a functional correlate of sarcomere addition in series [13,23,27]. Compared to other eccentric training studies, this change in FL shows to be 6% above the mean fiber lengthening reported for the lower limbs in a systematic review[18]. However, this review article also showed a wide range for adaptation, ranging from a 3% non-significant decrease in FL[33] to a possible 33% increase in FL for the hamstrings muscles[34]. Since the review article showed small differences in training method and exercise load between these two studies, the ultrasound imaging method applied was discussed as a possible reason for measuring different values in FL changes. Compared to MRI methods, 2D ultrasound imaging appears to be less objective and reliable[35]. To our knowledge, Suskens and collegues[21] published the first study using MRI-based mDTI and observed a 14% (2 cm) increase in FL after eccentric resistance training. While Suskens and collegues [21] trained the lower extremities, our study appears to be the first to demonstrate architectural changes of the shoulder after eccentric training using mDTI. However, plausible changes in FL were observed in this study, which are also reflected in the observed changes in FV. In terms of baseline fiber length and FV, the demonstrated results appear to be comparable to anatomical studies indicating a wide range of mean fiber length between 2.8 and 8.3 cm[36,37] and 6.6 cm for the infraspinatus muscle[37]. Since the population, measurement method, and number of fascicles measured can lead to incomparability between studies[36], this study appears to show results within the range for typical structural shape and adaption capacity.

### Limitations

There are several limitations that should be noted. First, this is a preliminary study without a control group and resulting in missing or bad data. However, this study was conducted to demonstrate the feasibility of training and diagnostics and to show the potential results for physically active men to have a basis for interpretation for future studies. Second, the sample size limited the statistical power and therefore some variables showed some trends but did not reach significance regardless of the analysis methods performed. Therefore, analysis revealed low sample size for the independent analysis for active stretching tests and MRI measurements and results have to be interpreted with caution. Third, the eccentric external rotation exercises used in this study may not account for strength training-induced changes in muscle architecture to the same extent as abduction exercises[29,38,39]. However, because of the widespread deficit in internal rotation of the glenohumeral joints as a risk factor for injury[40,41], rotational exercises have been used to directly target the major functional limitations of most subjects. Furthermore, the training with an isokinetic machine was very specific, whereas several subjects showed irritations in the torque-angle curves during training and testing. Therefore, it is possible that free-weight eccentric resistance training may produce at least the same effects as isokinetic training in such a population. Fourth, the stretching tests may not be comparable to other studies that primarily used physical therapists[29,38,39] to determine ROM of the glenohumeral joints. However, as most studies determine submaximal ROM without reaching an individual maximum stretch pain and with low standardization in how ROMmax is defined[17], this test is able to show stretching behavior in a standardized manner until a predefined torque or angle of motion is reached. However, the comparisons of curve morphology using SPM1d show an exploratory approach as there is no study to compare to. Last, differences in anatomical and diffusion-weighted MRI scan parameter settings led to difficulties in image registration and resampling. Because of these difficulties, four subjects were excluded from the infraspinatus analysis due to a proximally restricted field of view. In addition, the stopping criteria for fiber tracking resulted in interpolations and different tract densities between subjects. However, DTI represents the diffusion of tissue water molecules to calculate fiber length based on reconstructed tensors. Therefore, DTI results should be interpreted with caution.

## Conclusion

Following this study, it was shown that eccentric training can induce improvements in functional and structural measures of the shoulder external rotator cuff muscles. To our best knowledge, this is the first study using this experimental setting to determine baseline to posttest changes following eccentric strength training. Therefore, a major aim of this study was to evaluate the feasibility of the methods. In terms of feasibility, this study demonstrated that the eccentric intervention was able to confirm hypotheses regarding strength and muscle fiber length. Regarding the test methods, the e-function used to extrapolate a passive stretch torque-angle curve showed plausible results and was therefore considered feasible, as was DTI, which showed comparable results to the other studies mentioned in the Discussion section. Automatic segmentation approaches can be considered to speed up mDTI analysis. It remains to be proven whether these effects are still significant when compared to conventional shoulder prevention strategies in a randomized controlled trial. Furthermore, it is unknown whether this training program would produce similar results for a chronic high-impact shoulder for a sample of overhead athletes.

## Acknowledgments

We are grateful to all the scientists, support staff, and recruited individuals for their participation in this study. We would also like to thank Frank Yeh for the software support from DSI Studio.

## Supporting information

### Data availability

The dataset is available on a public repository on https://doi.org/10.6084/m9.figshare.24161040.v1

